# Symptom clusters in Covid19: A potential clinical prediction tool from the COVID Symptom study app

**DOI:** 10.1101/2020.06.12.20129056

**Authors:** Carole H. Sudre, Karla A. Lee, Mary Ni Lochlainn, Thomas Varsavsky, Benjamin Murray, Mark S. Graham, Cristina Menni, Marc Modat, Ruth C E Bowyer, Long H. Nguyen, David A. Drew, Amit D. Joshi, Wenjie Ma, Chuan-Guo Guo, Chun-Han Lo, Sajaysurya Ganesh, Abubakar Buwe, Joan Capdevila Pujol, Julien Lavigne du Cadet, Alessia Visconti, Maxim B Freidin, Julia S. El-Sayed Moustafa, Mario Falchi, Richard Davies, Maria F. Gomez, Tove Fall, M. Jorge Cardoso, Jonathan Wolf, Paul W. Franks, Andrew T. Chan, Tim D. Spector, Claire J Steves, Sébastien Ourselin

**Affiliations:** School of Biomedical Engineering & Imaging Sciences, King’s College London, Westminster Bridge Road, SE17EH London, UK; Department of Twin Research and Genetic Epidemiology, King’s College London, Westminster Bridge Road, SE17EH London, UK; Clinical and Translational Epidemiology Unit, Massachusetts General Hospital, MA, USA; Zoe Global Limited,164 Westminster Bridge Road, London SE1 7RW, UK; Lund University Diabetes Centre, Department of Clinical Sciences, Malmö, Sweden

**Author notes:** **Corresponding author(s):** Carole H Sudre and Sebastien Ourselin. These authors contributed equally to the work (Carole H. Sudre, Karla A. Lee, Mary Ni Lochlainn, Claire J Steves, Sébastien Ourselin).

## Abstract

As no one symptom can predict disease severity or the need for dedicated medical support in COVID-19, we asked if documenting symptom time series over the first few days informs outcome. Unsupervised time series clustering over symptom presentation was performed on data collected from a training dataset of completed cases enlisted early from the COVID Symptom Study Smartphone application, yielding six distinct symptom presentations. Clustering was validated on an independent replication dataset between May 1-May 28^th^, 2020. Using the first 5 days of symptom logging, the ROC-AUC of need for respiratory support was 78.8%, substantially outperforming personal characteristics alone (ROC-AUC 69.5%). Such an approach could be used to monitor at-risk patients and predict medical resource requirements days before they are required.

**One sentence summary:** Longitudinal clustering of symptoms can predict the need for respiratory support in severe COVID-19.

---

During the spread of the coronavirus disease 2019 (COVID-19) pandemic, the strain on healthcare systems has been felt globally and varying strategies for appropriate use of limited medical resource have been proposed (*1, 2*). However, heterogeneity in disease and presentation is evident, and the ability to predict required medical support ahead of time is limited. In this work, we sought to develop a clinical tool based on the time series of early development of COVID-19 that could be predictive of the need for high-level care in individuals more likely to seek medical help.

The COVID Symptom Study is a unique prospective population-based study collecting daily reports of symptoms from millions of users. The smartphone app offers a guided interface to report a range of baseline demographic information and comorbidities (as previously reported (*3, 4*)) and was developed by Zoe Global Limited with input from clinicians and scientists from King’s College London and Massachusetts General Hospital. With continued use, participants provide daily updates on symptoms, information on health care visits, COVID-19 testing results, and whether they are seeking medical support, including the level of intervention and related outcomes. Case reports have highlighted that COVID-19 infected individuals may present with different symptoms (*5*–*7*). We hypothesized that longitudinal symptoms reported during the illness would cluster into distinct subtypes with differing clinical needs and that we could use this information to create a predictive tool for medical support that could be used for resource planning and improvement of COVID 19 patient monitoring.

In order to study the time-series of symptom occurrence for the most severe cases for which respiratory support may be needed, clusters of longitudinally reported symptoms were obtained from an unsupervised clustering analysis (*8*) (see Methods). For our training dataset, we used data obtained from 1653 users of the app with persistent symptoms and regular logging, from disease onset until hospitalization or beginning of recovery, for which the data inclusion cut-off was the 30^th^ April 2020. An independent replication set was created using separate individuals fitting the criteria with a disease peak from the 30^th^ April to the 28^th^ May 2020. Patient selection is detailed online associated with a flow diagram (Sup Fig S1). The training sample for this analysis comprised 1,653 participants, of whom 383 reported at least one hospital visit and 107 reported respiratory support (defined as ventilation or supplementary oxygen). The independent replication sample consisted of 1,047 participants of which 207 reported a visit to hospital and 59 received respiratory support. Of participants in the independent replication set, 87.8% were from the United Kingdom, 7.5% were from the United States and 4.7% were from Sweden.

Prediction of the final cluster into which a participant would fall based on a short reporting period was assessed through tabulation of confusion matrices and weighted precision and recall. A predictive system focused on the need for respiratory support (supplemental oxygen or ventilation) was then built featuring the inferred cluster, the aggregated sum of symptoms and features of individual characteristics using five days of symptom reporting. Both clustering and predictive models were applied to the independent replication set of 1047 individuals.

Over the whole set of 2700 selected subjects, a number of demographic and health parameters were associated with higher risk for respiratory support requirement with the following odds ratios (OR) and 95% confidence intervals (95% CI): body mass index (BMI) in m/kg^2^ 1.05 per unit increase (95% CI [1.03 ;1.08], p<0.0005), older age OR 1.02, 95%CI: [1.01 ; 1.03], p=0.003), chronic lung disease (OR 2.72, 95%CI: [1.90 ; 3.90], p<0.0005), frailty as assessed by PRISMA7 questionnaire (*9*)(OR 5.98, 95%CI: [2.96 ; 12.10], p<0.0005) and a suggestive association with male sex (OR 1.49, 95%CI: [1.04 ; 2.13], p=0.029), respectively.

Unsupervised time series clustering (*5*) over the training set enabled us to distinguish six different clusters of symptom presentation. To visualize how clusters differed we used the reported average occurrence of a symptom on each day for the median duration (**Figure 1 top**) and the associated Z-Score for occurrence for each cluster with reference to the average presentation of one of the 14 reported symptoms (**Figure 1 bottom**). Equivalent plots for the independent replication dataset are presented as supplementary material (Figure S3)

**Figure 1.**
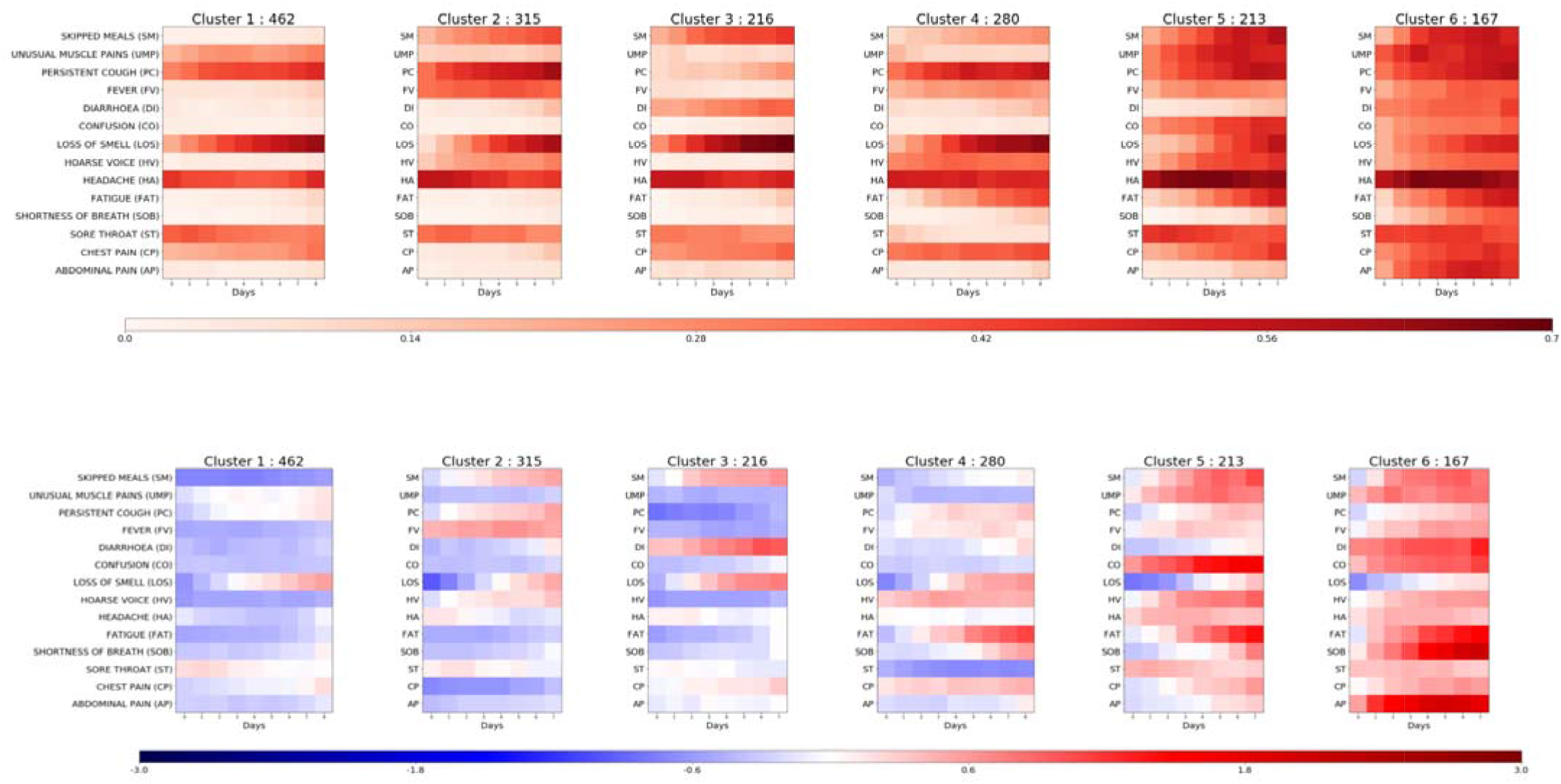
**Top:** Frequency of positive answers per symptoms across days for each cluster (darker = reported more frequently) and **Bottom**: associated Z-Score of presentation of symptoms over overall symptom distribution (red = reported more than average; blue = reported less than average). The clusters are ordered from left to right by rates of reported hospital visit with associated rates of respiratory support of 1.5%, 4.4%, 3.7%, 8.6%, 9.9% and 19.8% respectively.

Compared to Cluster 3 – 6, of which 8.6% - 19.8% required respiratory support, Cluster 1 and 2 represent milder forms of COVID-19 with 1.5% and 4.4% respectively, requiring respiratory support. These clusters showed predominantly upper respiratory tract symptoms and were distinguished from each other by the absence of muscle pain in Cluster 2 compared to Cluster 1, and slightly increased reports of skipped meals and fever in Cluster 2. Cluster 1 had notably lower mean age and BMI than the clusters containing patients with higher likelihood of requiring respiratory support (**Table 1**).

**Table 1.** Demographic details for the app users in each cluster, stratified by training/independent replication set membership. BMI = body mass index; Hosp=Hospital visit; RS = respiratory support. For continuous variables (age and BMI), results are given as mean (standard deviation); Frailty = PRISMA7 score >=3; Number is indicated with in parenthesis the origin of the participants (UK = United Kingdom, US = United States, SE = Sweden)

|  | Training 1653 (1621 – 32 - /) |  |  |  |  |  | Independent Replication 1047 (919 – 79 – 49) |  |  |  |  |  |
| --- | --- | --- | --- | --- | --- | --- | --- | --- | --- | --- | --- | --- |
|  | Cluster 1 | Cluster 2 | Cluster 3 | Cluster 4 | Cluster 5 | Cluster 6 | Cluster 1 | Cluster 2 | Cluster 3 | Cluster 4 | Cluster 5 | Cluster 6 |
| Number (UK – US – SE) | 462 (452 - 10 - /) | 315 (310 - 5 - /) | 216 (210 - 6 - /) | (273 - 7 - /) | 213 (211 - 2 - /) | 167 (165 - 2 - /) | 404 (348 - 35 - 21) | 199 (174 - 16 - 9) | 130 (115 - 9 - 6) | 102 (88 - 9 - 5) | 118 (111 - 4 - 3) | 94 (83 - 6 - 5) |
| Age | 41.1 (11.6) | 43.2 (12.1) | 41.0 (12.4) | 43.0 (11.9) | 43.7 (13.2) | 43.8 (12.2) | 44.2 (12.3) | 46.4 (14.1) | 45.9 (13.5) | 48.9 (14.5) | 48.1 (15.5) | 46.7 (15.8) |
| BMI | 26.9 (5.5) | 27.3 (5.7) | 26.9 (5.8) | 27.4 (5.9) | 28.0 (5.9) | 28.9 (6.3) | 27.1 (5.7) | 28.2 (5.9) | 28.6 (6.5) | 27.2 (5.3) | 28.7 (6.5) | 29.3 (6.2) |
| Male | 29.4% | 26.3% | 19.9% | 16.8% | 27.7% | 24.6% | 21.3% | 24.6% | 21.5% | 25.5% | 28.8% | 34.0% |
| Lung | 13.6% | 10.5% | 10.2% | 19.6% | 19.7% | 28.1% | 12.1% | 15.1% | 11.5% | 18.6% | 16.1% | 30.9% |
| Heart | 1.5% | 1.0% | 1.9% | 2.1% | 3.8% | 4.2% | 0.7% | 2.0% | 1.5% | 3.9% | 5.9% | 1.1% |
| Diabetes | 2.2% | 3.2% | 4.2% | 3.2% | 4.2% | 4.8% | 1.7% | 4.0% | 6.9% | 2.0% | 5.1% | 7.4% |
| Kidney | 0.4% | 0.6% | 0.5% | 1.1% | 0.9% | 1.8% | 0.5% | 0.5% | 0.0% | 0.0% | 1.7% | 2.1% |
| Frailty | 0.0% | 1.3% | 0.0% | 0.0% | 3.3% | 5.4% | 1.0% | 2.0% | 1.5% | 2.0% | 5.9% | 8.5% |
| Hosp | 16.0% | 17.5% | 23.6% | 24.6% | 27.2% | 45.5% | 11.6% | 13.1% | 13.8% | 24.5% | 37.3% | 50.0% |
| RS | 1.5% | 4.4% | 3.7% | 8.6% | 9.9% | 19.8% | 0.2% | 2.5% | 2.3% | 7.8% | 16.9% | 23.4% |

Cluster 3 shows stronger gastrointestinal symptoms in isolation (diarrhea, skipped meals) and a relatively reduced need for respiratory support, of 3.7%. However, the associated rate of hospital visit was high compared to cluster 1 and 2. Cluster 4, 5 and 6 included participants reporting more severe COVID-19 with 8.6%, 9.9% and 19.8% of individuals within these clusters requiring respiratory support, respectively. These three clusters represent distinct presentations, with Cluster 4 marked by the early presence of severe fatigue and the continuous presence of chest pain and persistent cough. In turn, individuals in cluster 5 reported confusion, skipped meals and severe fatigue. Finally, individuals in Cluster 6 reported more marked symptoms of respiratory distress including early onset of shortness of breath accompanied by chest pain. These respiratory symptoms were combined with significant abdominal pain, diarrhea and confusion when compared with other clusters. The proportion of frail people was higher in cluster 5 and 6 than in what we consider to be the *milder* clusters.

The ability to predict into which cluster a participant with COVID-19 will fall early in the disease process may enable the provision of adequate respiratory monitoring with pulse oximetry to at-risk patients. We used a confusion matrix analysis (as seen in **Figure 2**) and considered between two to nine days of recorded symptom data to perform the projection to different clusters. We found that after 5 days of reporting, despite 84.8% of the included samples presenting longer time-series in the training set, the error in projection was modest both in the training and the independent replication set. In this 6-class problem, the precision rose from 48.0% [45.9; 50.3] to 70.4% [68.4; 72.2] when moving from 2 to 5 days of data while the recall increased from 47.2% [45.1; 49.5] to 70.3% [68.4; 72.1]. Of note, when using 9 days of reported data, precision was 84.9% [83.5; 86.3] with a recall of 84.6% [83.2; 86.1].

**Figure 2.**
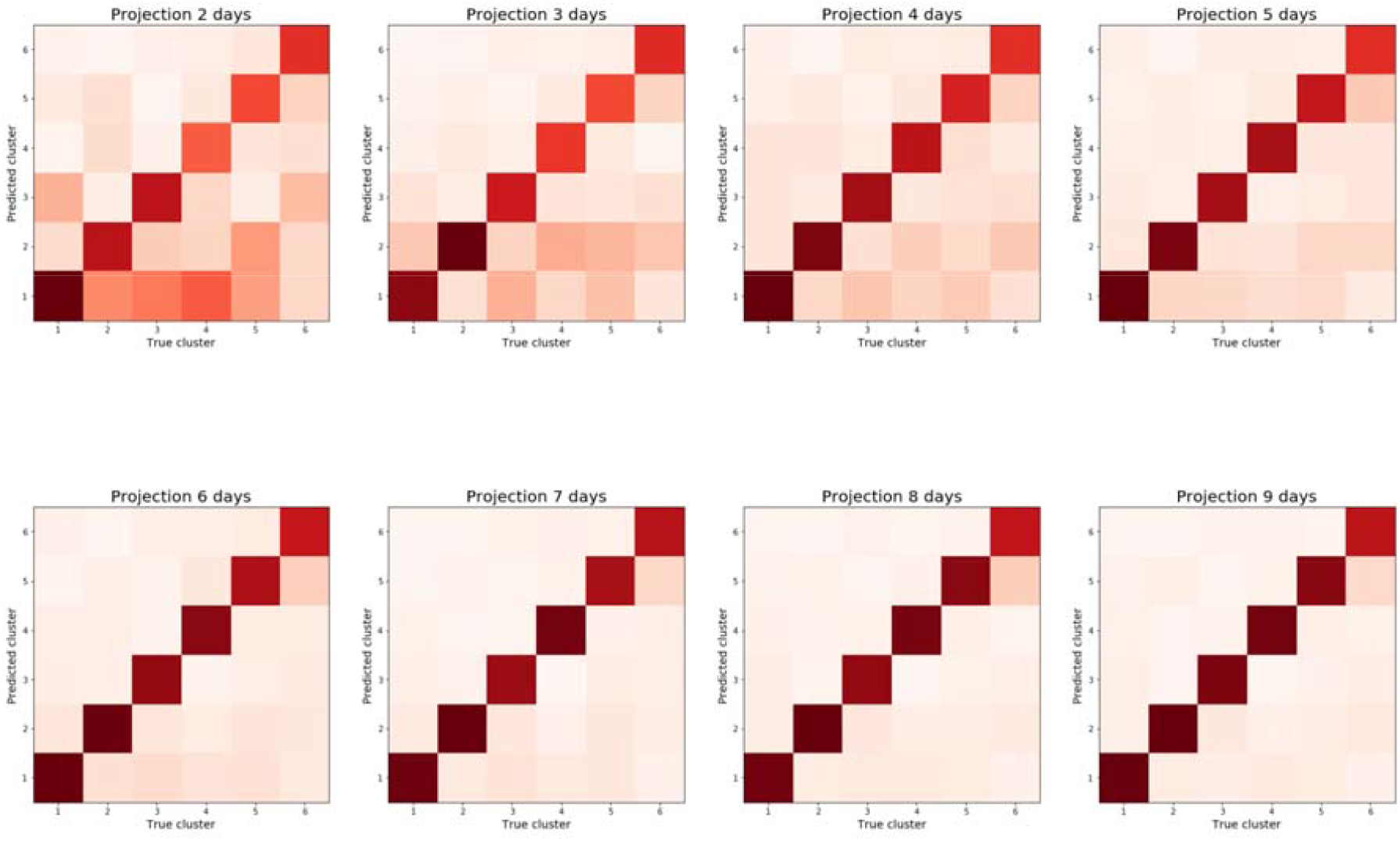
Confusion matrix showing cluster prediction using projections based on 2 to 9 days post onset of symptoms. By day 5 of COVID-19 the cluster in which a participant falls can be predicted with 72% weighted average precision.

**Figure 3:**
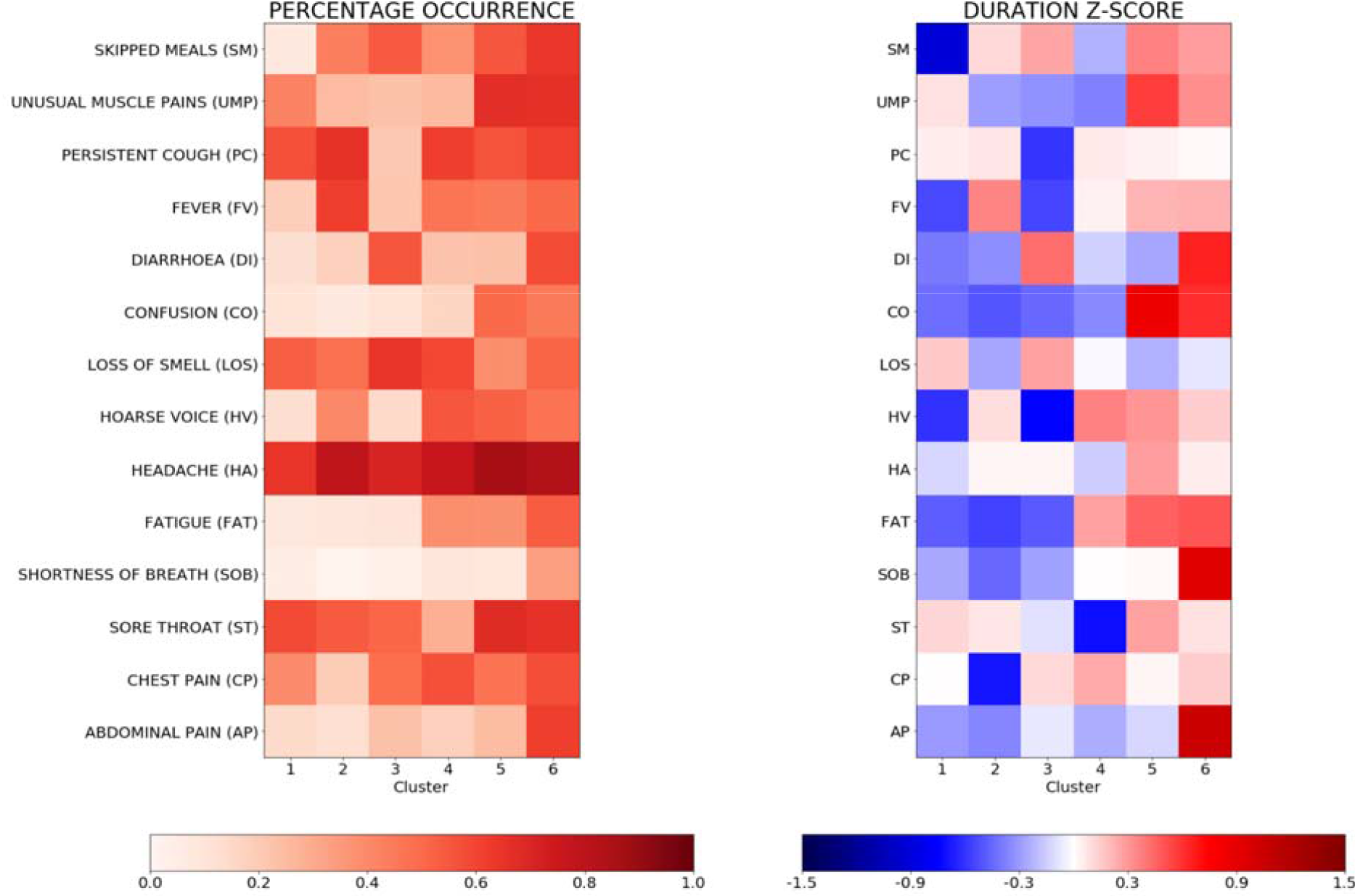
Left: Percentage of occurrence of symptoms at 5 days per cluster. Right: Z-Score in duration of symptom when occurring over the five first days.

The projections used to create the final clustering of the training set and those obtained from a reduced number of days were applied to the independent replication set and similar average patterns were observed: average precision and recall rose from (52.0 [49.4; 54.6], 51.8 [49.1; 54.3]) at two days to (72.4 [70.1; 74.6], 72.3 [70.1; 74.5]) at five days, and finally to (88.7 [87.1; 90.4], 88.7 [87.0; 90.3]) at nine days (see Figure S4).

At five days, it appeared that headache was the symptom most consistently reported across all clusters (see Figure 4), while severe fatigue appeared in those clusters with increased risk of requiring medical support. The duration of confusion was longer in more severe clusters while loss of smell or taste was reported over a longer duration in milder clusters.

While informative in their own right, we sought to develop a clinically useful tool using these clusters as a feature in a machine-learning-based system for predicting the need for respiratory support in COVID-19. Five days of reporting produced stable symptom clusters allowing for the construction of a predictive system that utilized data collected in the initial five days. The model used the predicted cluster (given 5 days reporting), the aggregated sum of symptoms up to and including that day, and personal characteristics including BMI, age, frailty (PRISMA7 score) and presence of comorbidities. The best model, trained with a 5-fold cross-validation and grid search hyper-parameters tuning, resulted in an area under the curve (AUC) of 78.8% [73.1; 84.2] on the independent replication set. In comparison, the demographic data alone led to an AUC of 69.5% [62.9; 74.5] where BMI, age and chronic lung disease were prominent features. Using the optimal Youden index derived from the training set (*10*), symptom information yielded a good recall (79.9% [60.3; 80.4]) with a false positive rate of 38.0% [35.3; 40.6], whereas, without symptoms, demographic and baseline health information led to a reduced recall (72.8 [63.2; 82.2]) with a larger false positive rate (46.5 [43.8; 49.0]), providing a clear argument for the inclusion of symptomatology alongside personal characteristics in prediction models for more severe forms of COVID-19.

Our study was limited by the use of self-reported information collected from individuals who used smartphone devices. Additionally, where individuals become too unwell to record their symptoms on the app later in the disease course, time series used in this work may not have accounted fully for the peak of the disease. To address this limitation, reporting-by-proxy was included on the app in late April 2020. National and regional differences in guidelines for hospital admissions and utilization of respiratory support exist, and given the multi-national nature of this study, must be acknowledged. In addition, our model cannot account for silent presentations such as cases of silent hypoxia reported in the literature (*11*). It must also be noted that due to the prospective design of the study and changes in population characteristics using the app, the independent replication set was observed to be older than the training set which may lead to slight over-estimation of severity in some younger individuals.

The ability to predict medical resource requirements days before they arise has significant clinical utility in this pandemic. If widely utilized, healthcare providers and managers could track large groups of patients and predict numbers requiring hospital care and respiratory support days ahead of these needs arising, allowing for staff, bed and intensive care planning. As a clinical tool, this approach could be implemented at local level, allowing patients to be monitored remotely by their primary healthcare teams with alert systems triggered when individuals demonstrate symptomatology associated with a high-risk cluster. Higher risk individuals could be targeted for increased care to ensure that they do not struggle to access advice when becoming more unwell. For instance, patients who fall into Cluster 5 or 6 at day 5 of the illness have a significant risk of hospitalization and respiratory support and may benefit from home pulse oximetry with daily phone calls from their general practice to ensure that hospital attendance occurs at the appropriate point in the course of their illness. Those in cluster 3 & 4 may also be at high risk, and benefit from proactive care, for example with glucose and electrolyte monitoring. A trigger system could be inbuilt as suggested in other initiatives (*12*), alerting these patients at high risk to seek medical attention at a point of specified predicted risk. Additionally, some patients and practitioners may be empowered by a clinical tool into which they could input longitudinal symptomatology and personal characteristics and receive personalised information on risk stratification.

## Data Availability

Data used in this study is available to bona fide researchers through UK Health Data Research using the following link https://healthdatagateway.org/detail/9b604483-9cdc-41b2-b82c-14ee3dd705f6

## Ethics

In the UK, the App Ethics has been approved by KCL ethics Committee REMAS ID 18210, review reference LRS-19/20-18210 and all subscribers provided consent. In Sweden, ethics approval for the study was provided by the central ethics committee (DNR 2020-01803).

## Funding

Zoe provided in kind support for all aspects of building, running and supporting the app and service to all users worldwide. Support for this study was provided by the NIHR-funded Biomedical Research Centre based at GSTT NHS Foundation Trust. This work was supported by the UK Research and Innovation London Medical Imaging & Artificial Intelligence Centre for Value Based Healthcare.

Investigators also received support from the Wellcome Trust, the MRC/BHF, Alzheimer’s Society, EU, NIHR, CDRF, and the NIHR-funded BioResource, Clinical Research Facility and BRC based at GSTT NHS Foundation Trust in partnership with KCL. ATC was supported in this work through a Stuart and Suzanne Steele MGH Research Scholar Award. CM is funded by the Chronic Disease Research Foundation and by the MRC AimHy project grant. LHN, DAD, ADJ, ADS, CG, WL are supported by the Massachusetts Consortium on Pathogen Readiness (MassCPR) and Mark and Lisa Schwartz. The work performed on the Swedish study is supported by grants from the Swedish Research Council, Swedish Heart-Lung Foundation and the Swedish Foundation for Strategic Research (LUDC-IRC 15-0067).

## Author contributions

C.H.S, K.L., M.N.L, C.J.S. and S.O conceived of and designed the experiments. C.H.S. analyzed the data. B.M., S.O., C.J.S., M.M, M.J.C., J.W., T.D.S., T.F., M.F.G, P.W.F., A.T.C., D.A.D and L.H.N. contributed reagents, materials and/or analysis tools. C.H.S, K.A.L and M.N.L wrote the manuscript. All authors revised the manuscript.

## Competing interests

Zoe Global Limited co-developed the app *pro bono* for non-commercial purposes. Investigators received support from the Wellcome Trust, the MRC/BHF, EU, NIHR, CDRF, and the NIHR-funded BioResource, Clinical Research Facility and BRC based at GSTT NHS Foundation Trust in partnership with KCL. RD, JW, JCP, AB, SG and JLduC work for Zoe Global Limited and TDS and PWF are consultants to Zoe Global Limited. LHN, DAD, PWF and ATC previously participated as investigators on a diet study unrelated to this work that was supported by Zoe Global Ltd.

